# Higher Expression of Denervation-responsive Genes is Negatively Associated with Muscle Volume and Performance Traits in the Study of Muscle, Mobility and Aging (SOMMA)

**DOI:** 10.1101/2023.11.04.23298090

**Authors:** Cole J. Lukasiewicz, Gregory J. Tranah, Daniel S. Evans, Paul M. Coen, Haley N. Barnes, Zhiguang Huo, Karyn A. Esser, Nancy E. Lane, Steven B. Kritchevsky, Anne B. Newman, Steven R. Cummings, Peggy M. Cawthon, Russell T. Hepple

## Abstract

With aging skeletal muscle fibers undergo repeating cycles of denervation and reinnervation. In approximately the 8^th^ decade of life reinnervation no longer keeps pace, resulting in the accumulation of persistently denervated muscle fibers that in turn cause an acceleration of muscle dysfunction. The significance of denervation in important clinical outcomes with aging is poorly studied. The Study of Muscle, Mobility and Aging (SOMMA) is a large cohort study with the primary objective to assess how aging muscle biology impacts clinically important traits. Using transcriptomics data from vastus lateralis muscle biopsies in 575 participants we have selected 49 denervation-responsive genes to provide insights to the burden of denervation in SOMMA, to test the hypothesis that greater expression of denervation-responsive genes negatively associates with SOMMA participant traits that included time to walk 400 meters, fitness (VO_2peak_), maximal mitochondrial respiration, muscle mass and volume, and leg muscle strength and power. Consistent with our hypothesis, increased transcript levels of: a calcium-dependent intercellular adhesion glycoprotein (CDH15), acetylcholine receptor subunits (Chrna1, Chrnd, Chrne), a glycoprotein promoting reinnervation (NCAM1), a transcription factor regulating aspects of muscle organization (RUNX1), and a sodium channel (SCN5A) were each negatively associated with at least 3 of these traits. VO_2peak_ and maximal respiration had the strongest negative associations with 15 and 19 denervation-responsive genes, respectively. In conclusion, the abundance of denervation-responsive gene transcripts is a significant determinant of muscle and mobility outcomes in aging humans, supporting the imperative to identify new treatment strategies to restore innervation in advanced age.

## Introduction

During the aging process skeletal muscle fibers undergo repeating cycles of denervation that are usually followed by successful reinnervation ^1^. This process profoundly remodels the spatial distribution of muscle fibers within the motor unit ^2^. Beyond this, the most significant impact of these recurring cycles of denervation-reinnervation is that eventually the rate of reinnervation cannot keep pace with denervation, resulting in the accumulation of muscle fibers that lack connection to a motor neuron ^3^. These persistently denervated muscle fibers progressively atrophy over time and can no longer contribute to force generation. For these reasons the accumulation of these persistently denervated fibers is an important contributor to both the reduction in muscle mass and contractile dysfunction with aging.

Rodent model studies find that alterations in neuromuscular junction morphology can be seen in some muscles relatively early in the lifespan (adulthood) ^4,5^, and this occurs well in advance of the accumulation of persistently denervated muscle fibers and the presentation of muscle atrophy with aging ^6^. Indeed, the accumulation of persistently denervated muscle fibers is a relatively late occurrence that can progress quite rapidly in advanced age (≥75 years old in humans), and impacts various skeletal muscle phenotypes ^7^ including mitochondrial function ^8,9^. Studies in rodent models of experimental denervation, e.g., sciatic nerve transection, have identified genes that respond (either increasing or decreasing depending upon the gene) to denervation and interestingly, many of these genes are also seen to change in aging muscle. For example, genes that regulate the stability of the acetylcholine receptor (AChR) cluster at the neuromuscular junction (e.g., muscle specific kinase [MuSK], AChR subunits, Agrin, rapsyn) are often seen to increase dramatically with surgical denervation ^10–13^, and many of these genes are also seen to increase in rodent ^14–16^ and human skeletal muscle ^8,17^ with aging. Hence, analysis of the expression levels of denervation-responsive genes can provide an indicator of the denervation burden in aging muscle. A significant question, however, concerns the importance of denervation in the decline in mobility with aging.

Previous studies examining how alterations in expression of denervation/neuromuscular junction genes are impacted with aging in humans have typically been small and have not had the power to establish how these alterations might relate to clinically important changes in physical function with aging. In this respect, the Study of Muscle, Mobility and Aging (SOMMA) is a large cohort study with the primary objective to assess how features of aging muscle biology relate to fitness, strength, muscle mass and other clinically relevant traits ^18^. Part of this study has involved acquiring muscle biopsies from the vastus lateralis muscle. Amongst the battery of measures performed with these biopsy specimens is whole transcriptome analysis using RNAseq. In this manuscript, using transcriptomics data from 575 participants we have selected 49 denervation-responsive genes to provide insights to the burden of denervation and test the hypothesis that expression levels of denervation-responsive genes are associated with walking speed, muscle mass, muscle strength/power, and fitness (VO_2peak_).

## Results

### 2.1 Participant Characteristics

Of the 879 participants who completed baseline measures, 591 had RNA sequencing completed and of these 575 had high quality sequencing and a complete set of covariate data for planned analyses. The characteristics of these 575 participants are provided in Table 1.

**Table 1.** Baseline characteristics of included SOMMA participants stratified by tertiles of VO2peak.

| Variable | Total<br>N | Total<br>Summary | Tertile<br>1 | Tertile<br>2 | Tertile<br>3 | P-Value | P-Trend |
| --- | --- | --- | --- | --- | --- | --- | --- |
|  |  |  | 653<br><=T1 <<br>1327 | 1327<br><=T2 <<br>1705 | 1705<br><=T3 <<br>3195 |  |  |
|  |  | (N=575) | (N=182) | (N=185) | (N=184) |  |  |
| **Clinic Site: Pittsburgh** | 575 | 263<br>(45.7) | 81<br>(44.5) | 86<br>(46.5) | 89<br>(48.4) | 0.760 | 0.459 |
| **Age, years** | 575 | 75.9 ±<br>4.5 | 77.3 ±<br>4.8 | 75.4 ±<br>4.4 | 74.9 ±<br>3.8 | <.001 | <.001 |
| **Sex: Female** | 575 | 319<br>(55.5) | 165<br>(90.7) | 121<br>(65.4) | 17<br>(9.2) | <.001 | <.001 |
| **Race: Non-Hispanic White** | 575 | 503<br>(87.5) | 158<br>(86.8) | 163<br>(88.1) | 165<br>(89.7) | 0.697 | 0.397 |
| **Height, m** | 575 | 1.7 ±<br>0.1 | 1.6 ±<br>0.1 | 1.7 ±<br>0.1 | 1.8 ±<br>0.1 | <.001 | <.001 |
| **Weight, kg** | 575 | 76.2 ±<br>15.5 | 66.3 ±<br>11.0 | 75.3 ±<br>13.3 | 86.8 ±<br>14.4 | <.001 | <.001 |
| **CHAMPS: Hours/week in**<br>**all exercise-related**<br>**activities** | 575 | 15.7 ±<br>11.4 | 13.7 ±<br>10.1 | 15.5 ±<br>9.7 | 18.5 ±<br>13.8 | 0.001 | <.001 |
| **Multimorbidity Count** | 575 |  |  |  |  | 0.394 | 0.200 |
| 0 Chronic conditions |  | 254<br>(44.2) | 79<br>(43.4) | 76<br>(41.1) | 90<br>(48.9) |  |  |
| 1 Chronic condition |  | 220<br>(38.3) | 71<br>(39.0) | 72<br>(38.9) | 70<br>(38.0) |  |  |
| 2+ Chronic conditions |  | 101<br>(17.6) | 32<br>(17.6) | 37<br>(20.0) | 24<br>(13.0) |  |  |
| **Maximal respiration**<br>**(pmol/(s\*mg))** | 519 | 61.5 ±<br>18.1 | 53.4 ±<br>13.5 | 62.5 ±<br>15.8 | 69.1 ±<br>20.9 | <.001 | <.001 |
| **Leg Strength: 1 repetition**<br>**maximum** | 559 | 177.9 ±<br>62.1 | 135.4 ±<br>37.4 | 171.8 ±<br>43.7 | 229.9 ±<br>61.0 | <.001 | <.001 |
| **400 m walk speed (m/s)** | 575 | 1.9 ±<br>0.2 | 1.0 ±<br>0.2 | 1.1 ±<br>0.2 | 1.1 ±<br>0.2 | <.001 | <.001 |
| **Total Thigh Fat Free**<br>**Muscle vol, L** | 557 | 9.1 ±<br>2.3 | 7.2 ±<br>1.2 | 8.8 ±<br>1.5 | 11.6 ±<br>1.6 | <.001 | <.001 |
| **D<sub>3</sub>CR muscle mass (kg)** | 551 | 22.6 ±<br>6.6 | 18.0 ±<br>3.6 | 21.6 ±<br>5.2 | 28.2 ±<br>6.1 | <.001 | <.001 |

Data shown as n (%), mean ± Standard Deviation. P-values are presented for variables across tertiles of VO<sub>2</sub> peak. P-values for continuous variables from ANOVA for normally distributed data, a Kruskal-Wallis test for skewed data. P for linear trend across categories was calculated with linear regression models for those normally distributed variables, a Jonckheere-Terpstra test for skewed data. P-values for categorical data from a chi-square test for homogeneity. P for trend was calculated with the Jonckheere-Terpstra test.

### 2.2 RNA (human Ensembl genes (ENSG)) detection

The mean, median, and SD of the PCR duplicate percent per sample was 59%, 56% and 9%, respectively (Supplementary Table 1). After PCR duplicates were removed, the number of aligned reads per sample was high (mean=69,117,209, median = 71,313,059, SD = 14,444,848, range = 12,853,785-102,724,183).

### 2.3 Associations with Age and other SOMMA Participant Traits

Fig 1 shows a volcano plot to illustrate the positive association between older age (at 5 y intervals) and greater expression of 13 denervation-responsive genes, where genes that satisfied an adjusted P-value of <0.05 are indicated in blue. We then characterized the association of SOMMA traits with the denervation-responsive genes reporting log base 2-fold changes reflecting the change in gene expression per one SD unit change in each trait. A summary of those genes that associated with at least 3 SOMMA traits is shown in a heatmap in Fig 2. Most of these associations were in the negative direction (an increase in transcript abundance associated with a lower value for a given trait); however, there were several transcripts for which we observed positive relationships between higher gene expression and a higher value for a given trait. This was most notable for UTRN which exhibited a positive association with 6 different traits. Notably, 3 of the genes with at least 3 significant negative associations encoded for structural components of the acetylcholine receptor cluster at the neuromuscular junction (CHRNA1, CHRND, CHRNE), one for a glycoprotein which helps attract neighboring axons to reinnervate denervated muscle fibers (NCAM1), and another encoded a sodium channel isoform found in muscle developmentally and which typically only appears in adult muscle following denervation (SCN5A). Turning to the specific trait associations, Fig 3 shows a Volcano plot of the genes that associated with walking speed. Of the 9 genes that reached an adjusted P-value of <0.05, 8 exhibited a negative association (RUNX1, THBS4, CHRNA1, CHRND, GADD45A, CDH15, NEFM and MYOG), whereas UTRN had a positive association with walking speed. VO_2peak_ (Fig 4) and maximal mitochondrial respiration (Fig 5) each demonstrated associations with the most denervation-responsive genes of all the traits examined and shared a common set of 11 genes for which there were negative associations (NEFM, CDH15, RUNX1, CHRNA1, CHRND, NCAM1, MUSK, CDK5R1, CAV3, SCN4A and SCN5A). Total thigh muscle volume demonstrated negative associations with 4 denervation-responsive genes, but positive associations with 9 denervation-responsive genes (Fig 6). Leg power demonstrated negative associations with 7 denervation-responsive genes (Fig 7), whereas leg strength demonstrated negative associations with 3 denervation-responsive genes (Fig 8). We found no significant associations between any of the denervation-responsive genes and D3Cr total body muscle mass. To provide an example of how biology independent of denervation may contribute to the associations observed, we also determined the association between UTRN and MHY7 (codes for slow myosin heavy chain in muscle fibers). As shown in Fig 9, there was a significant positive association between greater UTRN expression and MHY7 expression (r=0.301, P<0.001). A summary of the location and/or function of each of the genes that showed a significant negative or positive association with any of the participant traits is found in supplementary data (Supplementary Tables 1 and 2).

**Figure 1.**
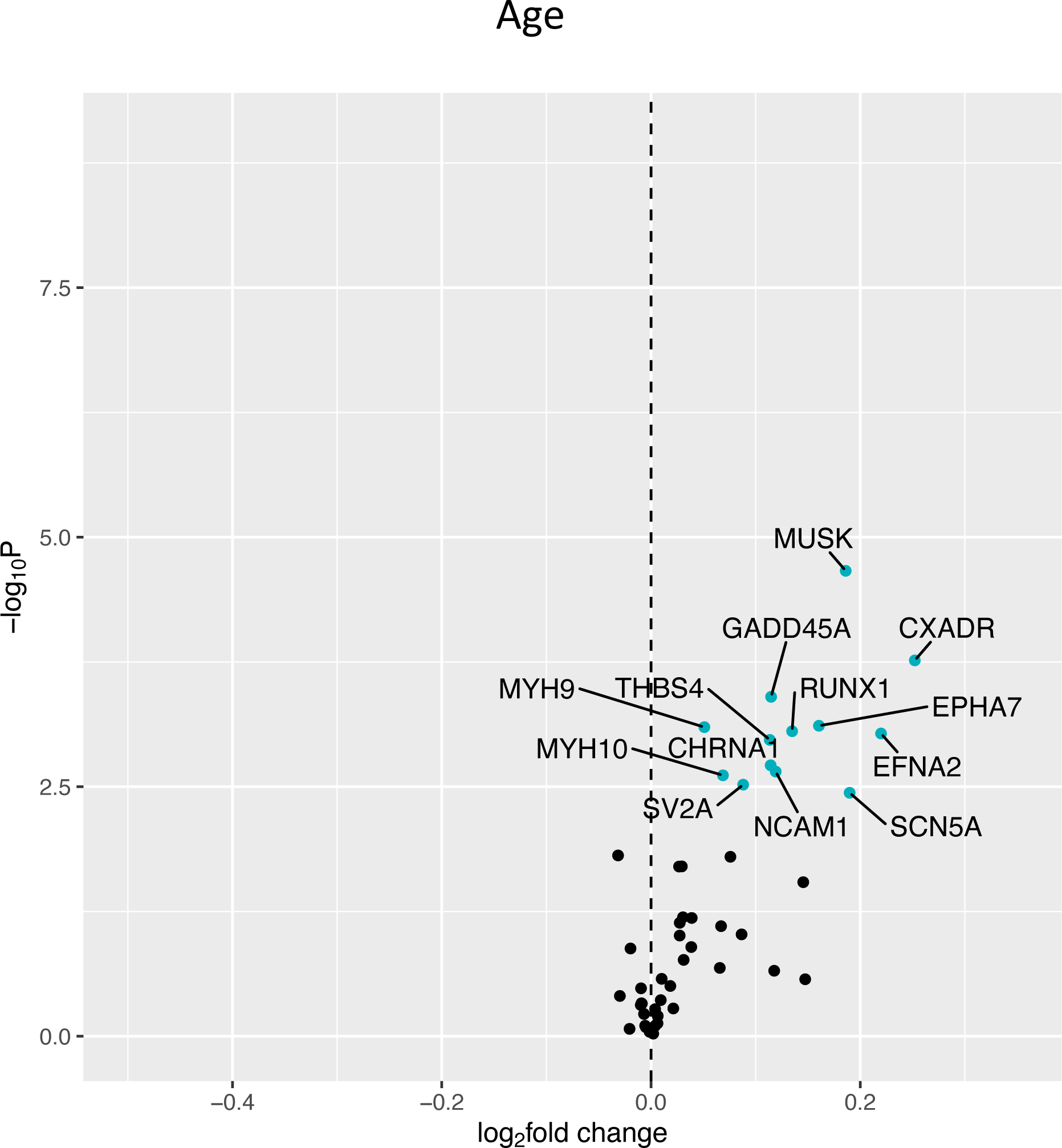
Associations with Age. Volcano plot capturing all statistically significant (p < 0.05 FDR adjusted) genes identified by our models: Each dot represents a gene; blue dots reached an adjusted P<0.05 threshold. Base model: gene expression∼age + sex + clinic site + race/ethnicity + body size + CHAMPS + multimorbidity count + sequencing batch.

**Figure 2.**
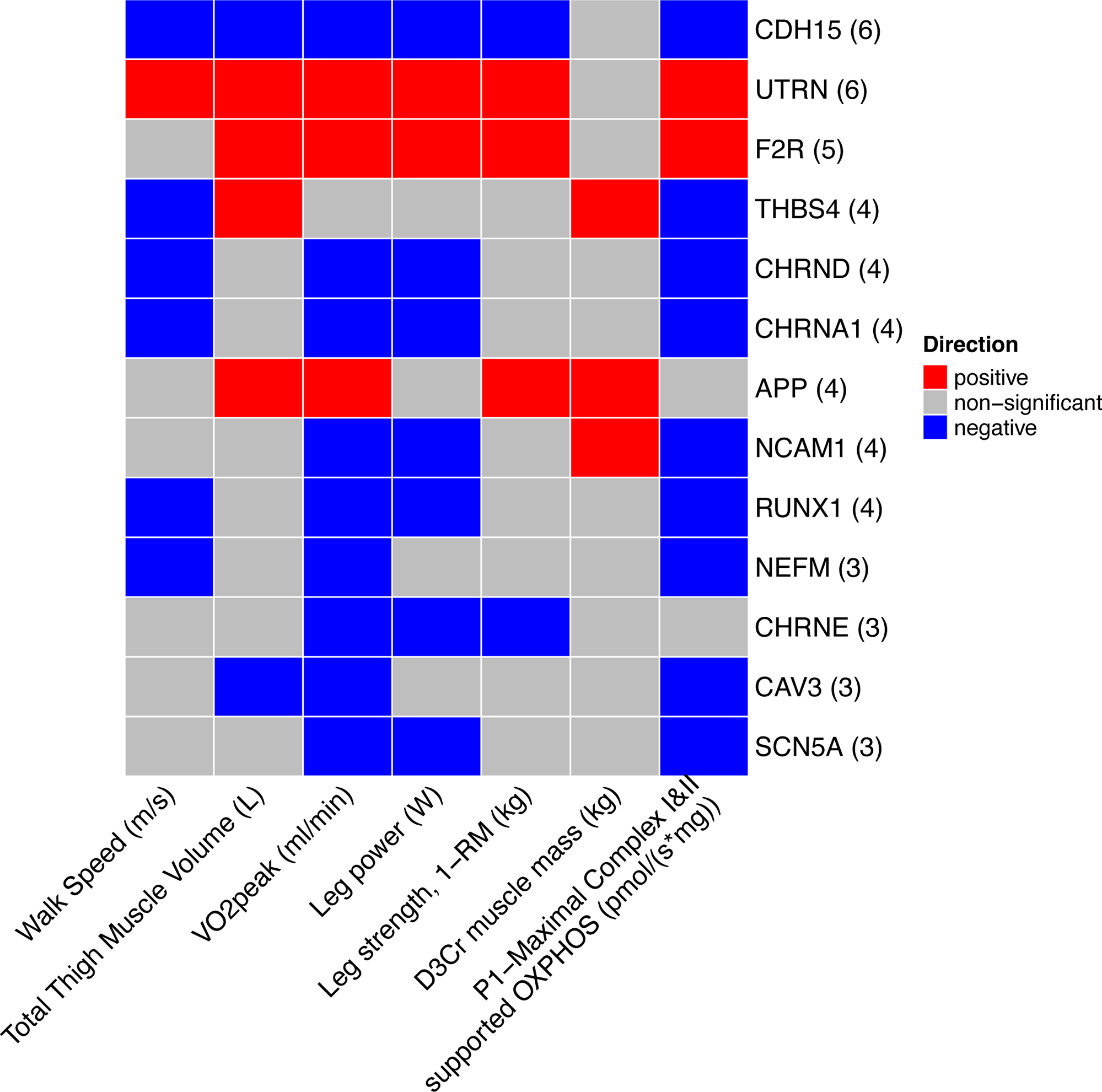
Significant associations with mitochondrial function, physical performance, and muscle mass measures. Heat map capturing all statistically significant (p < 0.05 FDR adjusted) genes identified by our models: each color represents positive (red) or negative (blue) associations.

**Figure 3.**
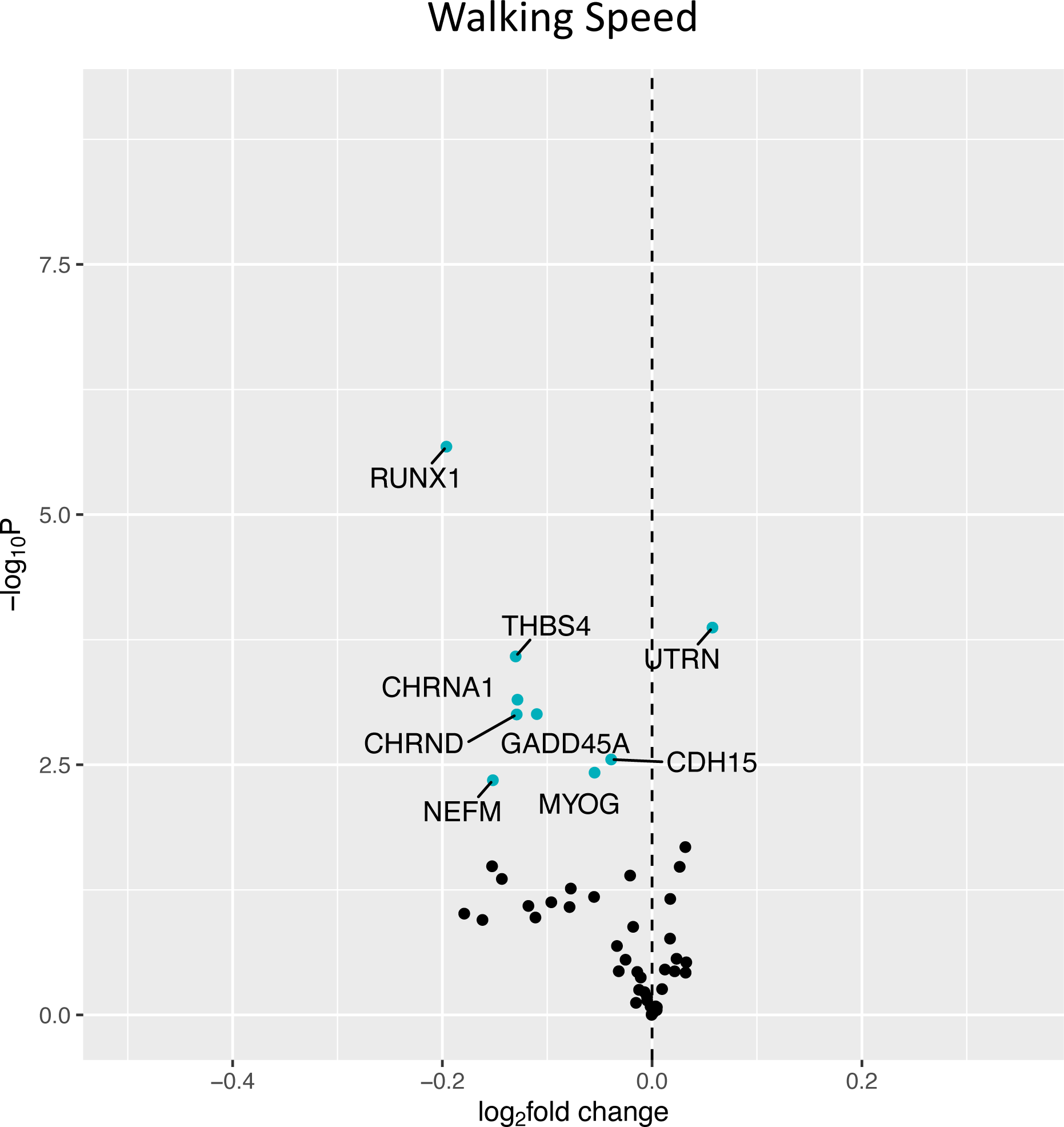
Associations with 400m Walk Speed. Volcano plot capturing all statistically significant (p < 0.05 FDR adjusted) genes identified by our models: Each dot represents a gene; blue dots reached an adjusted P<0.05 threshold. Base model: gene expression∼walk speed + age + sex + clinic site + race/ethnicity + body size + CHAMPS + multimorbidity count + sequencing batch.

**Figure 4.**
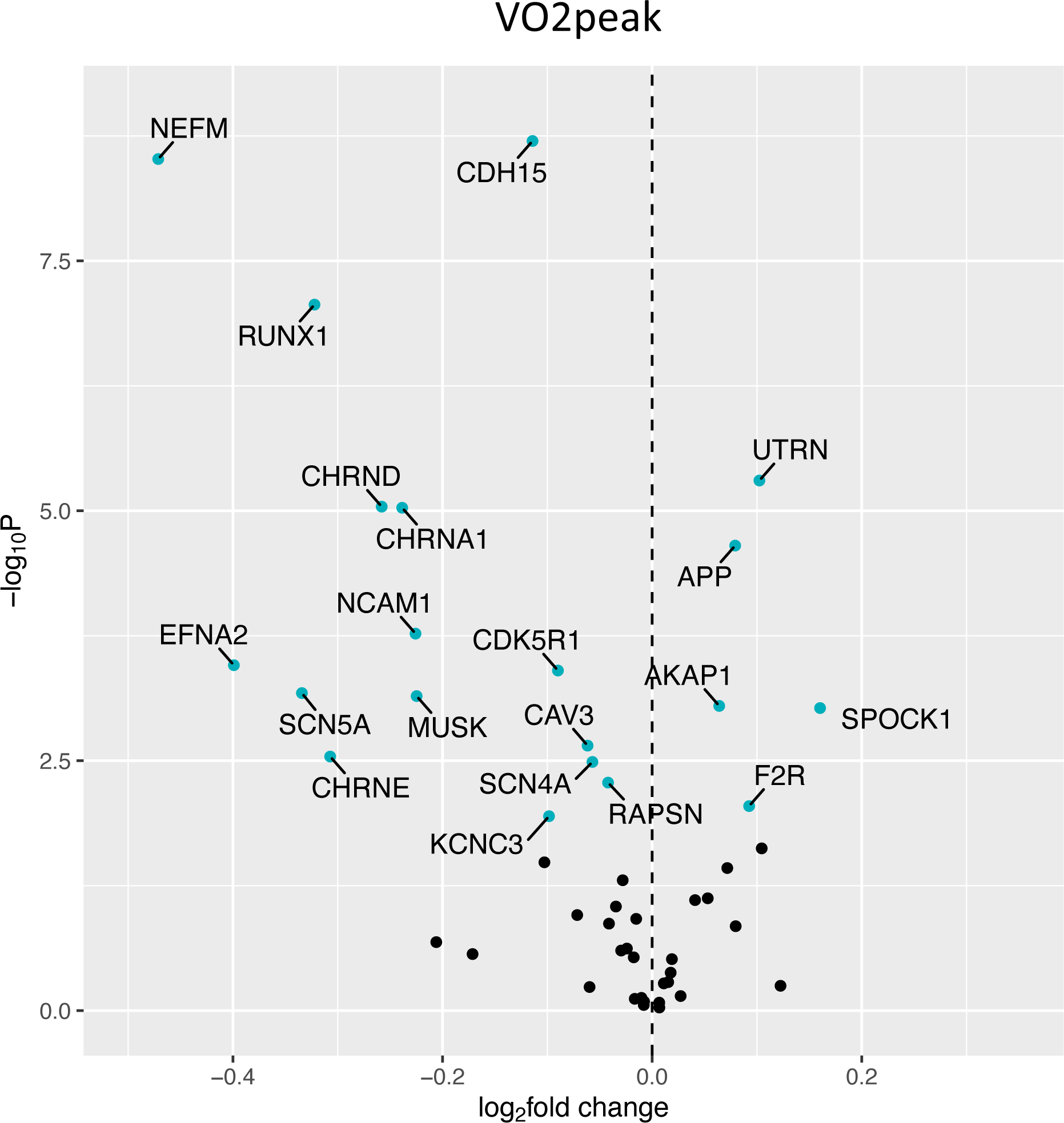
Associations with VO_2_ Peak. Volcano plot capturing all statistically significant (p < 0.05 FDR adjusted) genes identified by our models: Each dot represents a gene; blue dots reached an adjusted P<0.05 threshold. Base model: gene expression∼VO_2_ peak + age + sex + clinic site + race/ethnicity + body size + CHAMPS + multimorbidity count + sequencing batch.

**Figure 5.**
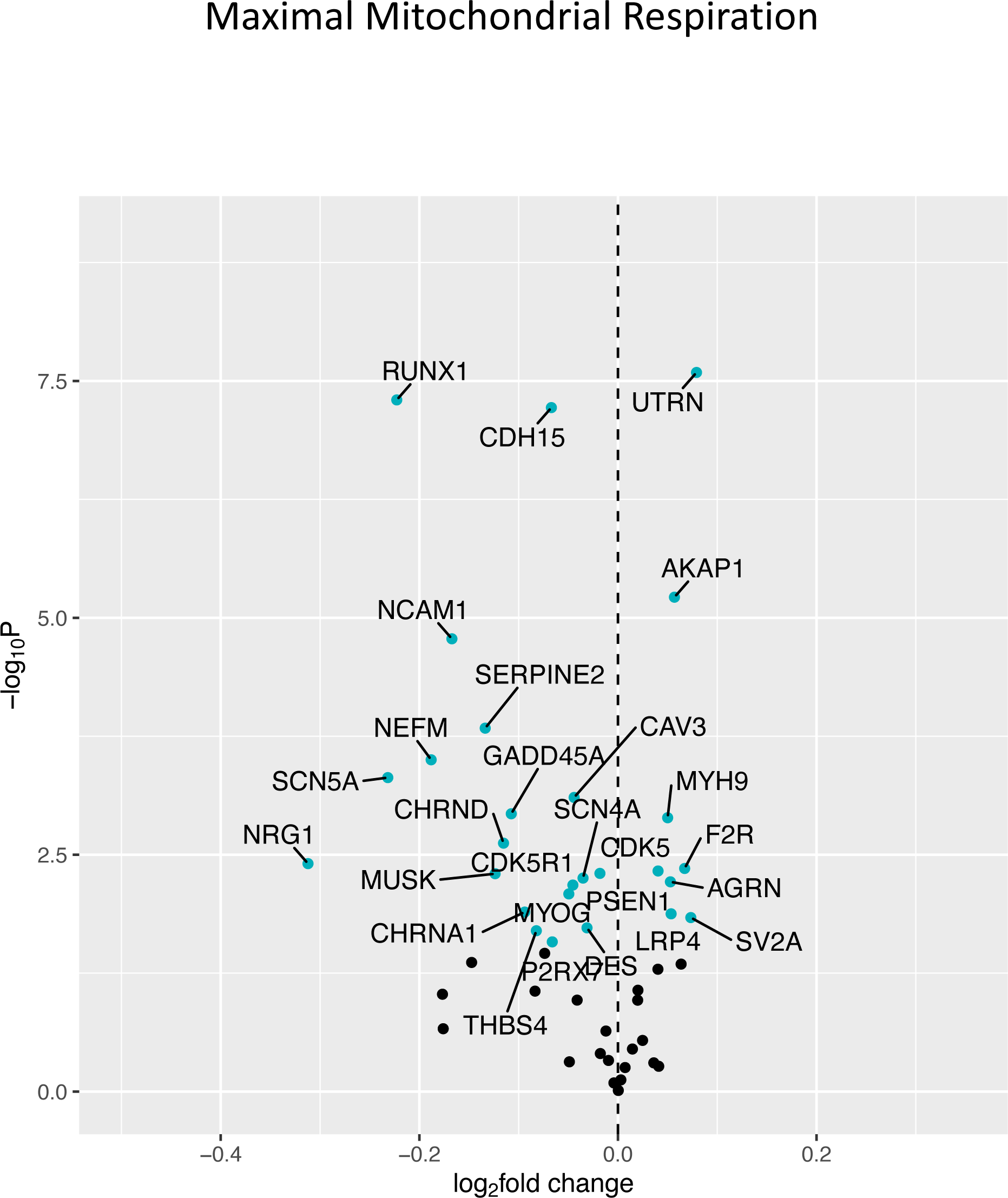
Associations with Maximal Mitochondrial Respiration. Volcano plot capturing all statistically significant (p < 0.05 FDR adjusted) genes identified by our models: Each dot represents a gene; blue dots reached an adjusted P<0.05 threshold. Base model: gene expression∼maximal respiration + age + sex + clinic site + race/ethnicity + body size + CHAMPS + multimorbidity count + sequencing batch.

**Figure 6.**
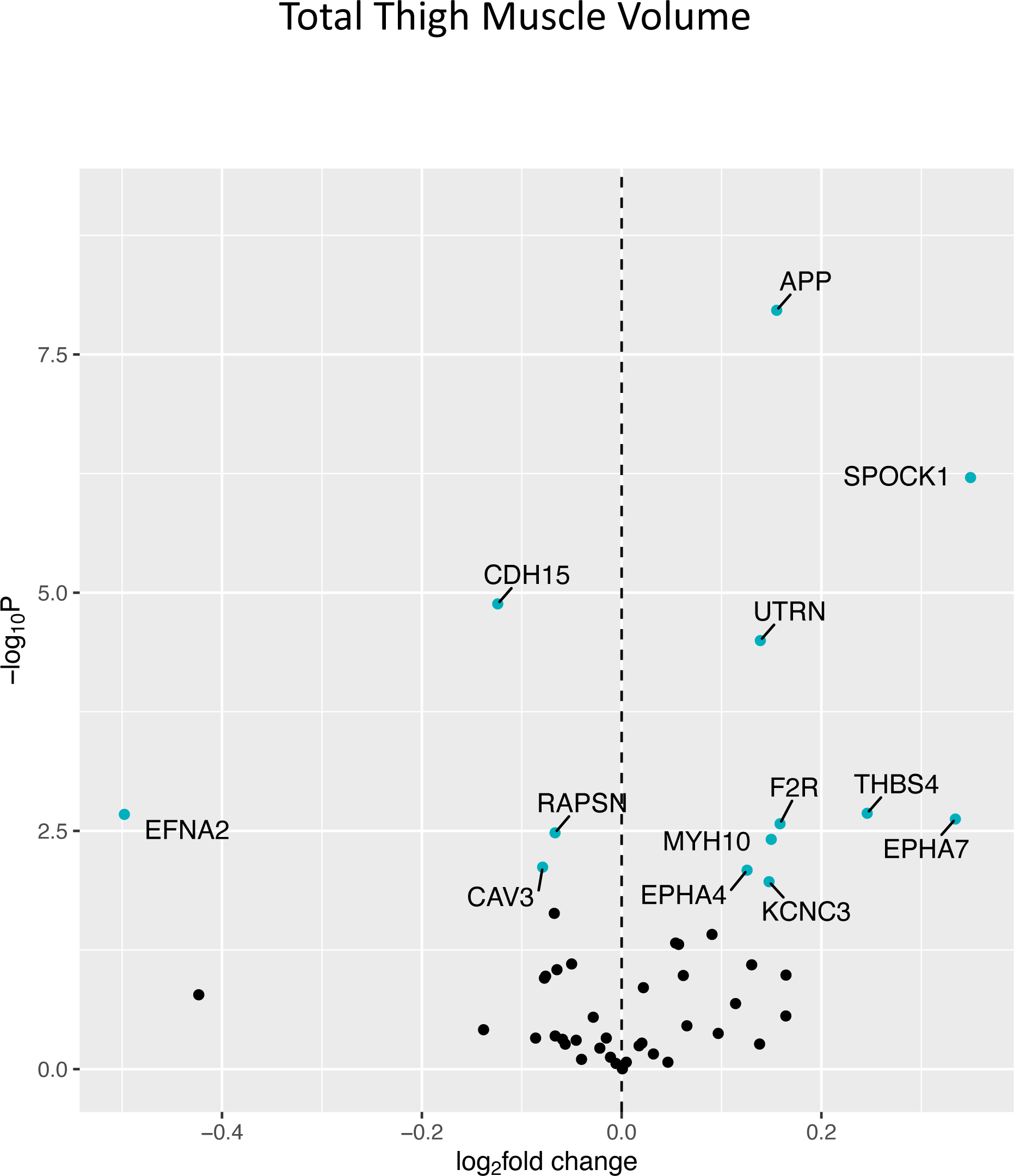
Associations with Total Thigh Muscle Volume. Volcano plot capturing all statistically significant (p < 0.05 FDR adjusted) genes identified by our models: Each dot represents a gene; blue dots reached an adjusted P<0.05 threshold. Base model: gene expression∼thigh muscle volume + age + sex + clinic site + race/ethnicity + body size + CHAMPS + multimorbidity count + sequencing batch.

**Figure 7.**
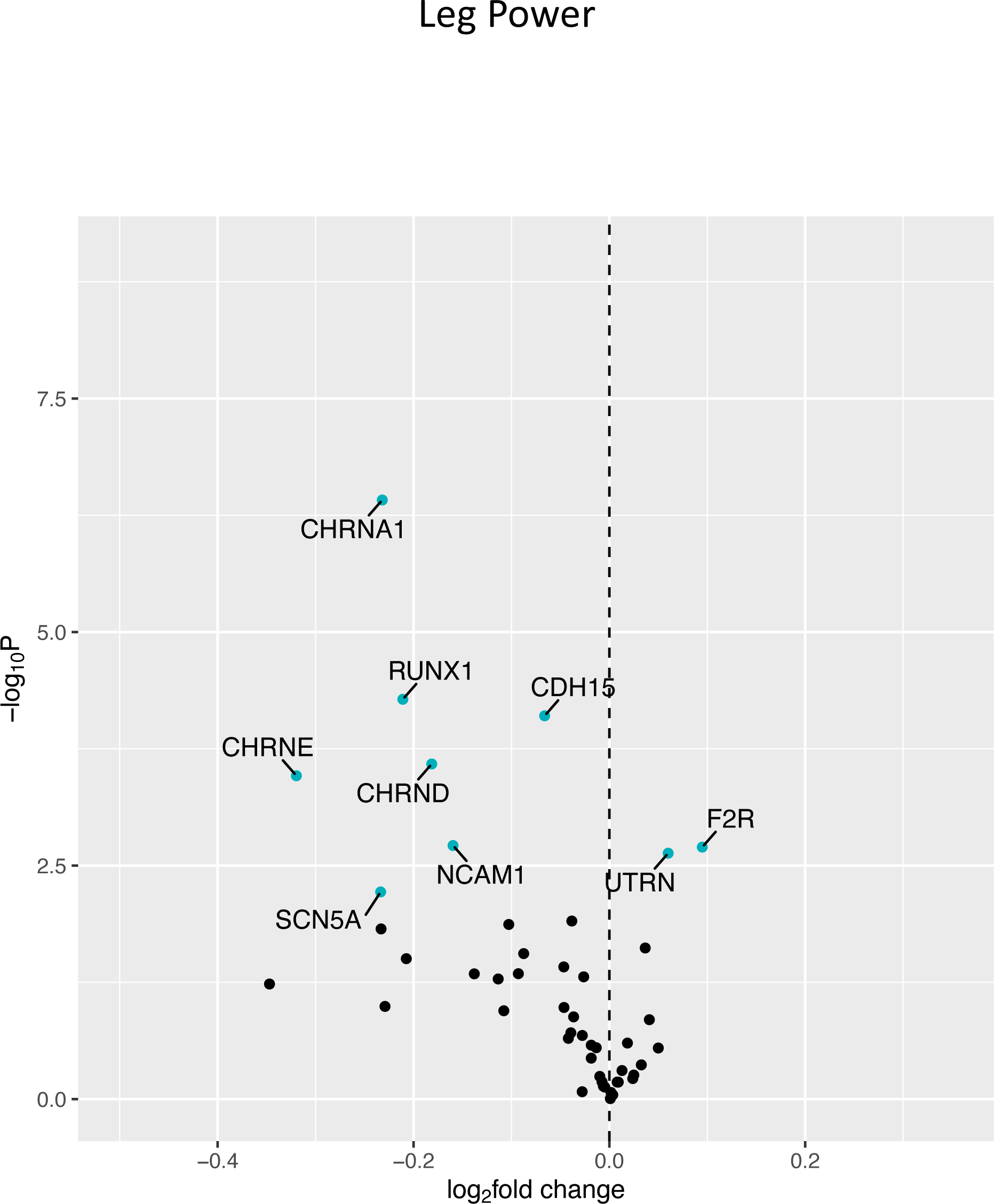
Associations with Leg Power. Volcano plot capturing all statistically significant (p < 0.05 FDR adjusted) genes identified by our models: Each dot represents a gene; blue dots reached an adjusted P<0.05 threshold. Base model: gene expression∼leg power + age + sex + clinic site + race/ethnicity + body size + CHAMPS + multimorbidity count + sequencing batch.

**Figure 8.**
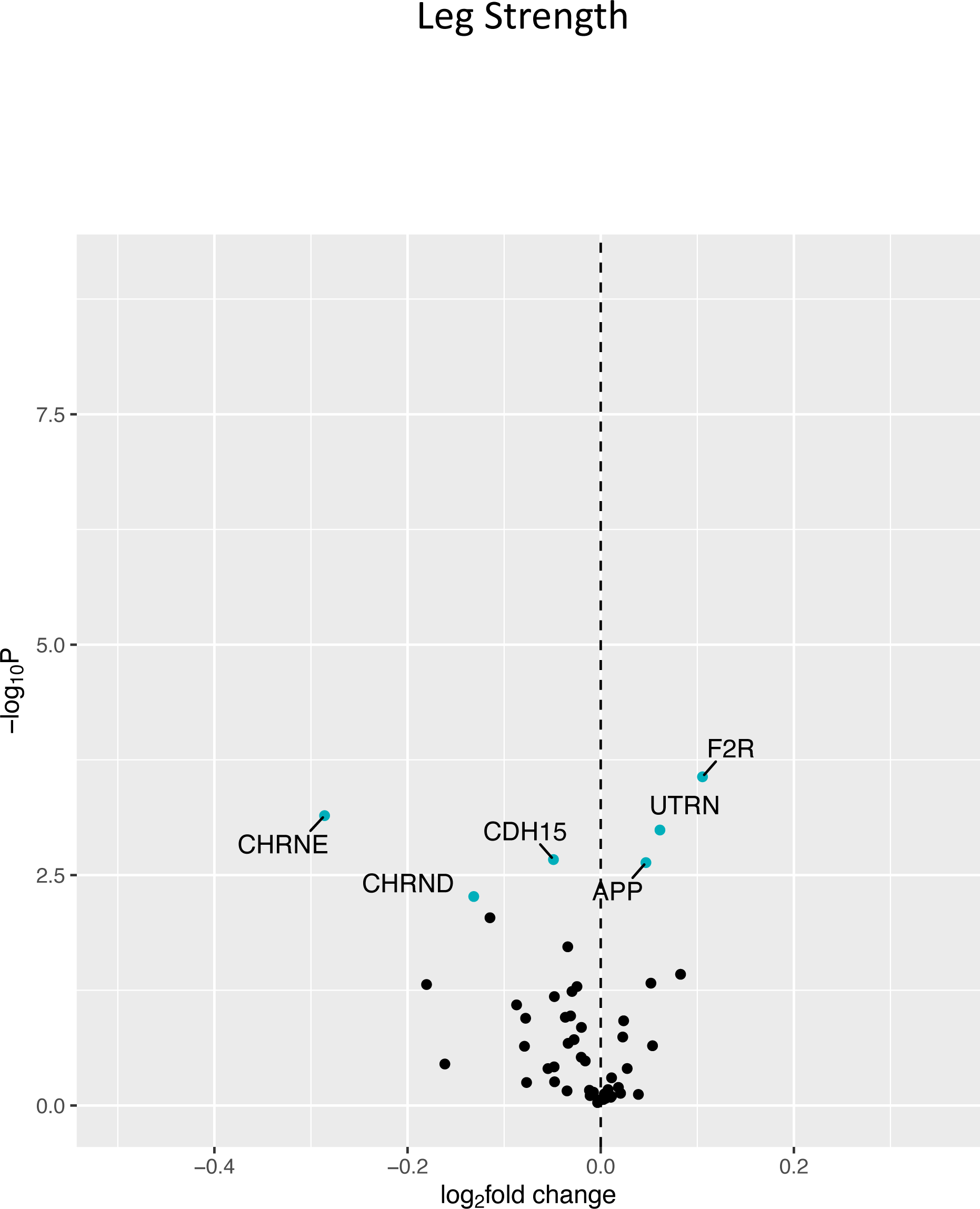
Associations with Leg Strength. Volcano plot capturing all statistically significant (p < 0.05 FDR adjusted) genes identified by our models: Each dot represents a gene; blue dots reached an adjusted P<0.05 threshold. Base model: gene expression∼leg strength + age + sex + clinic site + race/ethnicity + body size + CHAMPS + multimorbidity count + sequencing batch.

**Figure 9.**
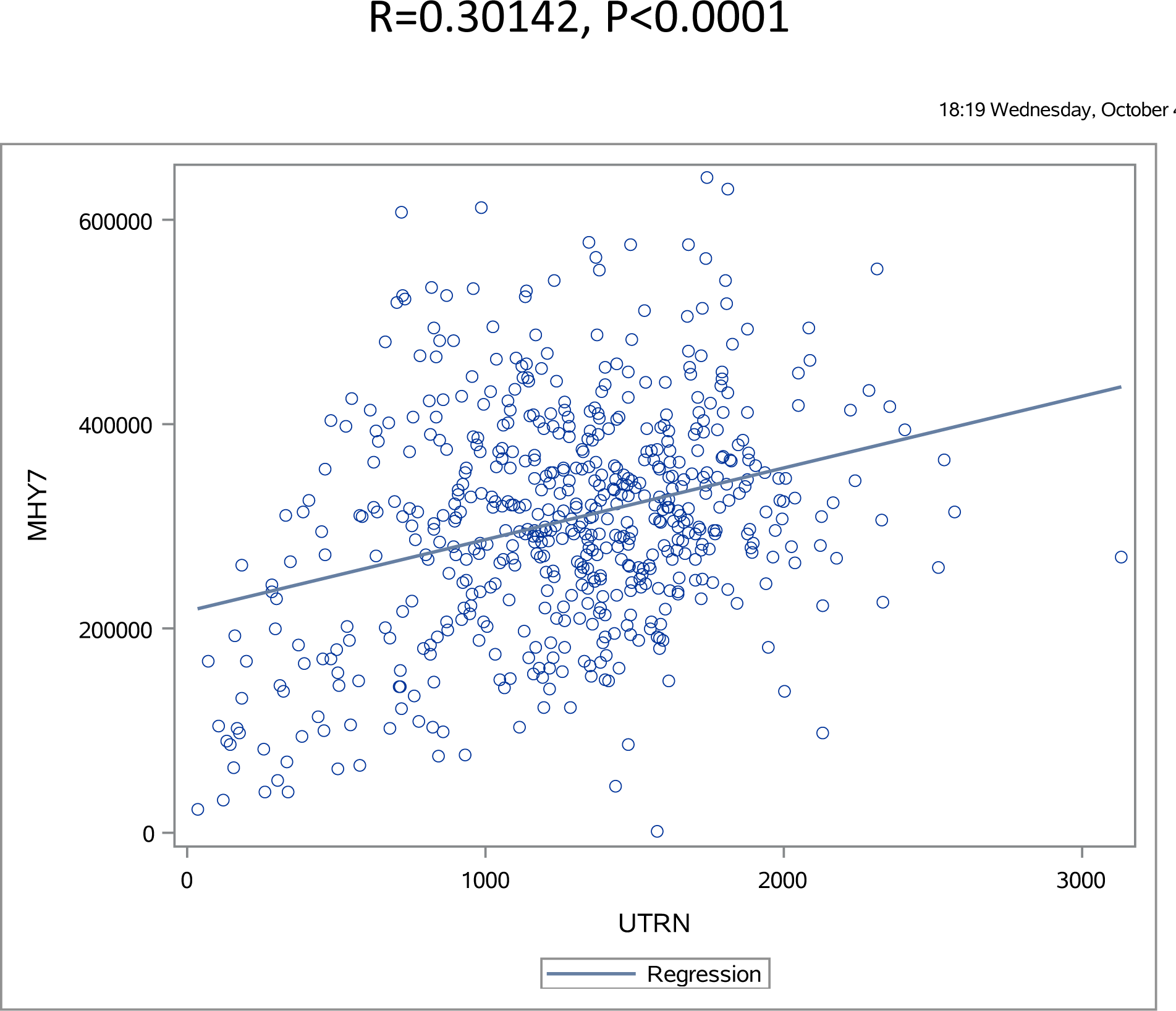
Association between UTRN and MYH7. Plot of UTRN expression in relation to MYH7 (slow myosin heavy chain isoform) expression in the SOMMA cohort.

## Discussion

The association of denervation with clinically relevant indices of strength, physical performance and muscle size with aging is poorly studied. To address this issue, we examined denervation-responsive transcript abundance using whole transcriptomic analyses of muscle biopsies from 575 healthy men and women ≥70 y old in the Study of Muscle, Mobility and Aging (SOMMA) and used sensitivity analyses to address how this associated with SOMMA participant traits that included walking speed, VO_2peak_, maximal mitochondrial respiration, muscle strength, muscle power, D3Cr total body muscle mass, and total thigh muscle volume. We hypothesized that greater abundance of denervation-responsive transcripts would associate positively with aging, but negatively with traits such as walking speed. Consistent with our hypothesis, higher expression of 13 denervation-responsive genes associated positively with increasing age. Furthermore, elevated expression of denervation-responsive genes was negatively associated with various functional and muscle size indices. For example, 400m walking speed was negatively associated with increased transcript abundance for genes encoding a calcium-dependent intercellular adhesion glycoprotein (CDH15), acetylcholine receptor subunits (CHRNA1, CHRND), denervation atrophy (GADD45A), and a transcription factor regulating aspects of muscle organization (RUNX1). Maximal mitochondrial respiration was negatively associated with increased transcript abundance for genes encoding CDH15, CHRNA1, CHRND, a glycoprotein that helps facilitate reinnervation (NCAM1), RUNX1, and a sodium channel (SCN5A). Similarly, leg power was negatively associated with elevated expression of genes encoding CDH15, CHRNA1, CHRND, CHRNE, another acetylcholine receptor subunit (CHRNE), NCAM1, RUNX1, and SCN5A. Collectively, our results are consistent with denervation playing an important role in driving declines in mobility and skeletal muscle function with aging in humans. In doing so, our findings underscore the importance of identifying strategies to maintain innervation of muscle fibers with aging.

Since the pioneering work of the Eccles which showed that switching the nerve supply between prototypical fast twitch versus slow twitch muscles made the fast muscles slower and the slow muscles faster ^19^, there has been an abundance of studies demonstrating the influence of muscle fiber innervation and muscle stimulation patterning on muscle phenotypes ^20–22^. It is thus not surprising that denervation of muscle fibers with aging is thought to play a very important role in driving hallmark features of aging muscle, including atrophy and fiber type shift ^1^. Although impaired neuromuscular junction transmission is considered a plausible contributor to impaired muscle function in aging humans ^23^, our understanding of how denervation relates to clinically important changes with aging, such as walking speed, is poorly studied. What we do know comes largely from smaller scale studies. For example, Piasecki and colleagues showed previously that older men with maintained muscle cross-sectional area had larger motor unit potentials than younger men, whereas older men with low muscle cross-sectional area did not. This finding was interpreted to indicate that the ability to reinnervate denervated muscle fibers, and thereby expand surviving motor units to compensate for those lost with aging, was essential to prevent muscle loss with aging ^24^. Similarly, our group have previously shown that motor unit number estimates are higher ^25^, and indices of denervation lower ^26^, in high functioning octogenarians. While these studies are generally supportive of an important role for denervation in muscle mass and mobility with aging, there is a clear need for additional study using a larger number of participants representing a range of age and function. In this respect, the Study of Muscle, Mobility and Aging (SOMMA) is a large cohort study whose primary objective is to address how skeletal muscle biology relates to mobility in 879 healthy men and women between the ages of 70 and 94 y ^18^.

In the current study we mined whole transcriptome RNAseq data from vastus lateralis muscle biopsies of 575 men and women (≥70 y old) who are SOMMA participants to determine how denervation-responsive genes associate with clinically important physical traits, such as walking speed. In this regard, there are many studies in the literature which have used rodent models to identify sets of genes that change in response to experimental denervation (e.g., sciatic nerve transection) ^10–12^, including recent work that takes this down to single nucleus resolution ^27^. In selecting the denervation-responsive genes for analysis in our current manuscript, we consulted these prior works, as well as reviews that address key components and signals involved in the mammalian neuromuscular junction (e.g., the agrin-Muscle Specific Kinase [MuSK] pathway, acetylcholine receptor [AChR] subunits, etc.) ^28,29^, and a prior study that specifically addressed the impact of aging on the skeletal muscle transcriptome where denervation genes were highlighted amongst those with the most significant changes with aging ^14^. It is important to note that several of the 49 denervation-responsive genes that we selected are also known to change in other contexts. As such, although all 49 of the genes selected are well-established as being responsive to denervation, there could be multiple processes influencing the expression of a given gene in aging muscle. It is likely that this pleiotropy of gene function and context accounts for the positive associations seen between some genes and SOMMA traits. As one example, amongst the denervation-responsive genes that we selected is UTRN, which codes for the protein utrophin. Utrophin is enriched at the neuromuscular junction and is known to increase in response to experimental denervation ^14^. However, utrophin is also enriched in oxidative versus glycolytic muscle fibers ^30^. Thus, the associations that we see in our dataset between elevated UTRN and higher walking speed, higher VO_2peak_, etc (which are directionally opposite to many other denervation-responsive genes) may reflect the influence of type I fiber abundance rather than denervation. Consistent with this idea, we observed a significant correlation between UTRN expression and expression of the gene encoding the myosin isoform found in type I muscle fibers (MHY7) in our dataset.

Notwithstanding the point above about some genes have functions beyond denervation, consistent with our hypothesis, we observed that increased expression of many of the selected denervation-responsive genes were negatively associated with clinically relevant measures in SOMMA participants. The clinical measures associated with the most denervation-responsive genes were VO_2peak_ (15 genes) and maximal mitochondrial respiration (19 genes), where 12 of the genes were common between these traits. The high degree of correspondence between VO_2peak_ and maximal mitochondrial respiration is not unexpected given the association of these traits in the SOMMA cohort ^31^ and in the Baltimore Longitudinal Study of Aging ^32^. What may be more surprising is that maximal mitochondrial respiration was associated with transcriptomic indices of denervation burden. In this respect, denervation has been shown to induce the release of fatty acid hydroperoxides (a form of reactive oxygen species [ROS]) from the mitochondrial outer membrane and this is catalyzed by the enzyme cytoplasmic phospholipase A2 (cPLA2) ^33^. Furthermore, pharmacological inhibition of cPLA2 using an agent called arachidonyl trifluoromethyl ketone (AACOCF3) normalized mitochondrial ROS from denervated muscle ^33^. Our group has used this strategy in aging humans and found that at ages associated with muscle morphological and transcriptional changes indicative of denervation cPLA2 inhibition using AACOCF3 reduced ROS emission in skeletal muscle mitochondria from elderly men ^8^ and women ^9^. This finding suggests that denervation is amongst the factors contributing to mitochondrial changes in aging skeletal muscle. Further to the influence of denervation on mitochondria, there is also a marked decrease in mitochondrial enzymes with denervation ^20^ secondary to the removal of mitochondria through mitophagy ^34^. As such, denervation is well-known to influence mitochondrial function and has also been implicated in altering some facets of mitochondrial function in aging human skeletal muscle previously. Our current findings add to this prior data in supporting the influence of denervation on mitochondrial function changes with aging.

Amongst the genes for which we observed significant negative associations with SOMMA traits was the transcription factor RUNX1. Interestingly, RUNX1 has been shown to be elevated in skeletal muscle of older patients with osteoporotic hip fracture and low muscle mass ^17^. Not only is RUNX1 elevated with denervation, but there is also a report observing a reduction in RUNX1 following resistance training in obese elderly participants and this was coincident with a reduction in the expression of the denervation-responsive glycoprotein NCAM1 at both the transcript level and protein level (NCAM protein) ^35^. The function of RUNX1 in the context of denervation was first described by Wang and colleagues where they showed that genetic knockout of RUNX1 led to an exacerbation of muscle impairment in the context of experimental denervation ^36^. They also identified that RUNX1 regulated the transcription of 29 genes that included ion channels, structural proteins, and signaling molecules. In this respect, a recent study identified RUNX1 as one of the transcriptions factors that can regulate SCN5A expression ^37^, noting that SCN5A was amongst the denervation-responsive genes that exhibited significant negative associations with VO_2peak_, maximal mitochondrial respiration, and leg power. Additional analyses of transcription factors related to the denervation-responsive genes in our dataset would be of interest in a future investigation.

## CONCLUSIONS

Our analysis of a set of denervation-responsive genes in 575 participants in SOMMA has revealed significant associations between increased expression of denervation-responsive genes and a diverse range of traits that include walking speed, VO_2peak_, maximal mitochondrial respiration, total thigh muscle volume, leg power and leg strength. Our results support and add to the evidence that denervation plays an important role in driving declines in various indices of function that contribute to the decline in mobility with aging.

## Methods

### Study Population

The Study of Muscle, Mobility and Aging (SOMMA) is a prospective cohort study of mobility in community-dwelling older adults. Participants for the current study were from the baseline cohort, enrolled between April 2019 and December 2021. SOMMA was conducted at 2 clinical sites: University of Pittsburgh (Pittsburgh, PA) and Wake Forest University School of Medicine (Winston-Salem, NC). Eligible participants were ≥70 years old at enrollment, had a body mass index (BMI) of 18–40 kg/m2, and were eligible for magnetic resonance (MR) imaging and a muscle tissue biopsy ^18^. Individuals with specific implants or on anticoagulation therapy were excluded due to ineligibility for MR imaging and muscle tissue biopsy, respectively. Individuals were further excluded if they had active cancer or were in the advanced stages of heart failure, renal failure on dialysis, dementia, or Parkinson’s disease. Participants must have been able to complete the 400 m walk; those who appeared as they might not be able to complete the 400 m walk at the in-person screening visit completed a short distance walk (4 meters) to ensure their walking speed as >=0.6m/s. Transportation to and from the clinic was provided to participants if needed for all study visits. The study protocol was approved by the Western Institutional Review Board Copernicus Group (WCG IRB; study number 20180764) and all participants provided written informed consent.

More information about SOMMA recruitment and study protocols can be found elsewhere ^18^. In brief, baseline testing occurred across 3 clinic visits that were completed up to within 8 weeks of each other. The average time between Day 1 and Day 3 testing was 42 days (6 wk). Day 1 included general clinic assessments (e.g., physical tests; 5 hours), Day 2 included magnetic resonance imaging and Cardiopulmonary Exercise Testing (MR and CPET, 2–3 hours), and Day 3 included fasting specimen and tissue collection (2 hours). There were 879 participants who completed Day 1 of baseline testing and had at least one primary SOMMA measure: CPET, MR imaging, or muscle tissue biopsy.

### Study Measures

#### Cardiorespiratory fitness (VO_2_ peak)

Cardiorespiratory fitness was measured using gold-standard VO_2_ peak from Cardiopulmonary Exercise Testing (CPET). A standardized CPET, using a modified Balke or manual protocol, was administered to participants to measure ventilatory gases, oxygen and carbon dioxide inhaled and exhaled during exercise ^38^. Participants who were excluded from the maximal effort symptom-limited peak test had acute electrocardiogram (ECG) abnormalities, uncontrolled blood pressure or history of myocardial infarction, unstable angina or angioplasty in the preceding 6 months. Testing for VO_2_ peak began at the participant’s preferred walking speed with incremental rate (0.5 mph) and/or slope (2.5%) increased in 2-minute stages until respiratory exchange ratio, ratio between VCO_2_ and VO_2_, was ≥1.05 and self-reported Borg Rating of Perceived Exertion (REF) was ≥17. Blood pressure, pulse oximetry, and ECG were monitored throughout exercise. VO_2_ peak was determined in the BREEZESUITE software (MGC Diagnostics, St. Paul, MN) as the highest 30-second average of VO2 (mL/min) achieved. The data were manually reviewed to ensure the VO_2_ peak was selected correctly for each participant.

### Other Measures

*P*articipants are asked to walk at their usual pace for 400 m from which walking speed was calculated. Whole-body D3Cr muscle mass was measured in participants using a D3-creatine dilution protocol as previously described ^39^. Knee extensor leg power was assessed using a Keiser Air 420 exercise machine in the same leg as the muscle biopsy. Resistance to test power was based on determination of the 1 repetition maximum leg extensor strength. Weight was assessed by balance beam or digital scales and height by wall-mounted stadiometers. An approximately 6-minute long MR scan was taken of the whole body to assess body composition including thigh muscle volume with image processing by AMRA Medical ^40^. The CHAMPS questionnaire ^41^ was used to assess specific types and the context of physical activities.

### Sample Preparation and Sequencing

#### Skeletal Muscle Biopsy Collection and Processing

Percutaneous biopsies were collected from the middle region of the musculus vastus lateralis under local anesthesia using a Bergstrom canula with suction (34). Following this, the specimen was blotted dry of blood and interstitial fluid and dissected free of any connective tissue and intermuscular fat. Approximately 20 mg of the biopsy specimen was placed into ice-cold BIOPS media (10 mM Ca–EGTA buffer, 0.1 M free calcium, 20 mM imidazole, 20 mM taurine, 50 mM potassium 2-[N-morpholino]-ethanesulfonic acid, 0.5 mM dithiothreitol, 6.56 mM MgCl2, 5.77 mM ATP, and 15 mM phosphocreatine [PCr], pH 7.1) for respirometry, as previously described (22). Myofiber bundles of approximately 2–3 mg were teased apart using a pair of sharp tweezers and a small Petri dish containing ice-cold BIOPS media. After mechanical preparation, myofiber bundles were chemically permeabilized for 30 minutes with saponin (2 mL of 50 μg/mL saponin in ice-cold BIOPS solution) placed on ice and a rocker (25 rpm). Myofiber bundles were washed twice (10 minutes each) with ice-cold MiR05 media (0.5 mM ethylenediaminetetraacetic acid, 3 mM MgCl2·6H2O, 60 mM K-lactobionate, 20 mM taurine, 10 mM KH2PO4, 20 mM N-2-hydroxyethylpiperazine-Nʹ-2-ethanesulfonic acid, 110 mM sucrose, and 1 g/L bovine serum albumin, pH 7.1) on an orbital shaker (25 rpm). The second wash in MiR05 contained blebbistatin (25 μM), a myosin II ATPase inhibitor, that was used to inhibit muscle contraction. Fiber bundle wet weight was determined immediately after permeabilization using an analytical balance (Mettler Toledo, Columbus, OH).

#### RNA Library Preparation and Sequencing

Total RNA from frozen human skeletal muscle samples (∼5 to 30mg) was prepared using Trizol solution (Invitrogen) according to the manufacturer’s direction in 2.0mL Eppendorf safe-lock tubes. Homogenization was performed using the Bullet Blender (NextAdvance, Raymertown NY USA) with an appropriate amount of stainless steel beads (autoclaved, 0.5∼2mm, NextAdvance, Raymertown NY USA) at 4°C on Setting 8 (Max is 12) in 30 second bouts. The homogenization step was repeated 5 times for a total of 3 minutes with at least 1 minute break between each bout. The removal of residual genomic DNA was performed by incubating the RNA sample with DNase (AM1907, Thermosci) plus RiboLock RNase inhibitor (EO0381, Thermisci) at 37°C for 25min in a heating block (400rpm). Cleanup of the RNA samples was done using the DNase inactivation reagent following instructions provided instructed by the manufacturer (AM1907, Thermosci). The RNA concentration and integrity were determined by using ThermoSci Nanodrop and Agilent Tapestation.

To prepare RNAseq library, polyA mRNA was isolated from about 250ng total RNA by using NEBNext Poly(A) mRNA magnetic isolation module (E7490L, NEB) and mRNA library constructed by using NEBNext Ultra II directional RNA library Pre Kit for Illumina (E7760L, NEB). Equal molarity of RNAseq libraries were pooled and sequenced on Illumina NovaSeq (2X150bp) to reach 80M reads per sample.

### Alignment and Quality Control

The reads from RNA-sequencing were aligned to the Genome Reference Consortium Human Build 38 (GRCh38) using the HISAT2 software ^42^. Duplicated aligned reads were further marked and excluded using the Picardtools software. Expression count data were obtained using the HTseq software (PMID 25260700). Genes with a total count of ≤ 20 across all samples were filtered out to remove non-expressed genes.

### Pathway Identification

For the analyses performed in this manuscript, we manually curated a list of 49 genes whose expression is altered in surgical denervation studies ^10–13^ and which have previously been shown to be enriched amongst the differentially expressed genes seen in aging rat skeletal muscle ^14^. These include genes involved in maintenance of the AChR cluster, genes encoding the 5 AChR subunits, genes promoting reinnervation, genes encoding different isoforms of sodium channels, and genes involved in the muscle atrophy pathway. In addition, we selected genes encoding 6 neurotrophins that regulate maintenance and sprouting of motoneurons ^43–45^, and genes encoding 6 neurotrophin receptors.

### Regression Models and Visualization

Expression levels of forty-nine denervation-responsive genes were analyzed. Gene expression associations with traits were identified using negative binomial regression models as implemented by DESeq2 ^46^ in R and adjusted for age, gender, clinic site, race/ethnicity, height, weight, hours per week in all exercise-related activities (CHAMPS), multimorbidity count category, and sequencing batch. DESeq2 uses a negative binomial generalized linear model for differential analysis and applies the Wald test for the significance of GLM coefficients. The Benjamini-Hochberg false discovery rate method was used for P-value adjustment. Genes were considered differentially expressed according to the significance criteria of FDR<0.05. In negative binomial models, traits were modeled using the number of standard deviations (SDs) from each trait’s mean.

Consequently, the reported log base 2-fold changes reflect the change in gene expression per one SD unit increase in each trait. Volcano plots were created to visualize the differential expression of RNAs (ENSGs) associated with functional measures. Heat maps were created to summarize significant ENSG associations across all analyzed traits.

## Statement relating to relevant ethics and integrity policies

The study protocol was approved by the Western Institutional Review Board Copernicus Group (WCG IRB; study number 20180764) and all participants provided written informed consent.

## Acknowledgments

For a full list of personnel who contributed to the SOMMA study, please see Cummings SR, Newman AB, Coen PM, Hepple RT, Collins R, Kennedy K, Danielson M, Peters K, Blackwell T, Johnson E, Mau T, Shankland EG, Lui LY, Patel S, Young D, Glynn NW, Strotmeyer ES, Esser KA, Marcinek DJ, Goodpaster BH, Kritchevsky S, Cawthon PM. The Study of Muscle, Mobility and Aging (SOMMA). A Unique Cohort Study about the Cellular Biology of Aging and Age-related Loss of Mobility. J Gerontol A Biol Sci Med Sci. 2023 Feb 9:glad052. doi: 10.1093/gerona/glad052. Epub ahead of print. PMID: 36754371.

## Authors’ contributions

SC, AN, SK, RTH, PMCa, KE and DE designed the study.

HB, ZH, DE, KAE, CL, RTH, GT conducted the main analyses and drafted the manuscript.

KAE, ZH, DE, HB conducted the genetic analyses.

RTH, KE, DE, ZH, NEL contributed to the interpretation of the results.

KAE, ZH contributed to data collection.

RTH, SC, PMCa, AN, NEL, SK, PMCo critically revised the manuscript.

All authors reviewed and approved the final version of the manuscript and KE, ZH, and HB had full access to the data in the study and accept responsibility to submit for publication.

## Data availability statement

All SOMMA data are publicly available via a web portal. Updated datasets are released approximately every 6 months https://sommaonline.ucsf.edu/.

## Supplementary information

**Supplemental Table 1:** Gene name, location and/or function for genes with which SOMMA traits had a negative association. Only gene associations that reached an adjusted P<0.05 are listed.

**Supplemental Table 2:** Gene name, location and/or function for genes with which SOMMA traits had a positive association. Only gene associations that reached an adjusted P<0.05 are listed.

**Supplemental Table 3:** RNAseq quality metrics for each sample, including: number of raw pairs, alignment rate, number of aligned reads, duplication rate, and number of aligned reads deduplicated.

## References

1 Hepple, R. T. & Rice, C. L. Innervation and neuromuscular control in ageing skeletal muscle. J Physiol 594, 1965–1978, doi:10.1113/JP270561 (2016).

2 Larsson, L. Motor units: remodeling in aged animals. J Gerontol A Biol Sci Med Sci 50 Spec No, 91–95 (1995).

3 Anagnostou, M. E. & Hepple, R. T. Mitochondrial Mechanisms of Neuromuscular Junction Degeneration with Aging. Cells 9, doi:10.3390/cells9010197 (2020).

4 Valdez, G. et al. Attenuation of age-related changes in mouse neuromuscular synapses by caloric restriction and exercise. Proc Natl Acad Sci U S A 107, 14863–14868, doi:1002220107 [pii] 10.1073/pnas.1002220107 (2010).

5 Burke, S. K., Fenton, A. I., Konokhova, Y. & Hepple, R. T. Variation in muscle and neuromuscular junction morphology between atrophy-resistant and atrophy-prone muscles supports failed re-innervation in aging muscle atrophy. Exp Gerontol 156, 111613, doi:10.1016/j.exger.2021.111613 (2021).

6 Deschenes, M. R., Roby, M. A., Eason, M. K. & Harris, M. B. Remodeling of the neuromuscular junction precedes sarcopenia related alterations in myofibers. Exp Gerontol 45, 389–393, doi:10.1016/j.exger.2010.03.007 (2010).

7 Rowan, S. L. et al. Denervation causes fiber atrophy and Myosin heavy chain co-expression in senescent skeletal muscle. PLoS One 7, e29082, doi:10.1371/journal.pone.0029082 (2012).

8 Spendiff, S. et al. Denervation drives mitochondrial dysfunction in skeletal muscle of octogenarians. J Physiol 594, 7361–7379, doi:10.1113/JP272487 (2016).

9 Sonjak, V. et al. Reduced Mitochondrial Content, Elevated Reactive Oxygen Species, and Modulation by Denervation in Skeletal Muscle of Prefrail or Frail Elderly Women. J Gerontol A Biol Sci Med Sci 74, 1887–1895, doi:10.1093/gerona/glz066 (2019).

10 Mugahid, D. A. et al. Proteomic and Transcriptomic Changes in Hibernating Grizzly Bears Reveal Metabolic and Signaling Pathways that Protect against Muscle Atrophy. Scientific reports 9, 19976, doi:10.1038/s41598-019-56007-8 (2019).

11 Macpherson, P. C., Farshi, P. & Goldman, D. Dach2-Hdac9 signaling regulates reinnervation of muscle endplates. Development 142, 4038–4048, doi:10.1242/dev.125674 (2015).

12 Soares, R. J. et al. Involvement of microRNAs in the regulation of muscle wasting during catabolic conditions. J Biol Chem 289, 21909–21925, doi:10.1074/jbc.M114.561845 (2014).

13 Ebert, S. M. et al. Stress-induced skeletal muscle Gadd45a expression reprograms myonuclei and causes muscle atrophy. J Biol Chem 287, 27290–27301, doi:10.1074/jbc.M112.374777 (2012).

14 Ibebunjo, C. et al. Genomic and proteomic profiling reveals reduced mitochondrial function and disruption of the neuromuscular junction driving rat sarcopenia. Mol Cell Biol 33, 194–212, doi:10.1128/MCB.01036-12 (2013).

15 Aare, S. et al. Failed reinnervation in aging skeletal muscle. Skeletal muscle 6, 29, doi:10.1186/s13395-016-0101-y (2016).

16 Barns, M. et al. Molecular analyses provide insight into mechanisms underlying sarcopenia and myofibre denervation in old skeletal muscles of mice. Int J Biochem Cell Biol 53, 174–185, doi:10.1016/j.biocel.2014.04.025 (2014).

17 Kang, Y. J., Yoo, J. I. & Baek, K. W. Differential gene expression profile by RNA sequencing study of elderly osteoporotic hip fracture patients with sarcopenia. J Orthop Translat 29, 10–18, doi:10.1016/j.jot.2021.04.009 (2021).

18 Cummings, S. R. et al. The Study of Muscle, Mobility and Aging (SOMMA). A Unique Cohort Study about the Cellular Biology of Aging and Age-related Loss of Mobility. J Gerontol A Biol Sci Med Sci, doi:10.1093/gerona/glad052 (2023).

19 Buller, A. J., Eccles, J. C. & Eccles, R. M. Interactions between motoneurones and muscles in respect of the characteristic speeds of their responses. J Physiol 150, 417–439 (1960).

20 Abruzzo, P. M. et al. Oxidative stress in the denervated muscle. Free radical research 44, 563–576, doi:10.3109/10715761003692487 (2010).

21 Ausoni, S., Gorza, L., Schiaffino, S., Gundersen, K. & Lomo, T. Expression of myosin heavy chain isoforms in stimulated fast and slow rat muscles. J Neurosci 10, 153–160 (1990).

22 Patterson, M. F., Stephenson, G. M. M. & Stephenson, D. G. Denervation produces different single fiber phenotypes in fast- and slow-twitch hindlimb muscles of the rat. AJP - Cell Physiology 291, 518–528 (2006).

23 Arnold, W. D. & Clark, B. C. Neuromuscular junction transmission failure in aging and sarcopenia: The nexus of the neurological and muscular systems. Ageing Res Rev 89, 101966, doi:10.1016/j.arr.2023.101966 (2023).

24 Piasecki, M. et al. Failure to expand the motor unit size to compensate for declining motor unit numbers distinguishes sarcopenic from non-sarcopenic older men. J Physiol 596, 1627–1637, doi:10.1113/JP275520 (2018).

25 Power, G. A. et al. Motor unit number and transmission stability in octogenarian world class athletes: Can age-related deficits be outrun? J Appl Physiol (1985) 121, 1013–1020, doi:10.1152/japplphysiol.00149.2016 (2016).

26 Sonjak, V. et al. Fidelity of muscle fibre reinnervation modulates ageing muscle impact in elderly women. J Physiol 597, 5009–5023, doi:10.1113/JP278261 (2019).

27 Lin, H. et al. Decoding the transcriptome of denervated muscle at single-nucleus resolution. Journal of cachexia, sarcopenia and muscle 13, 2102–2117, doi:10.1002/jcsm.13023 (2022).

28 Rudolf, R., Khan, M. M., Labeit, S. & Deschenes, M. R. Degeneration of Neuromuscular Junction in Age and Dystrophy. Frontiers in aging neuroscience 6, 99, doi:10.3389/fnagi.2014.00099 (2014).

29 Tintignac, L. A., Brenner, H. R. & Ruegg, M. A. Mechanisms Regulating Neuromuscular Junction Development and Function and Causes of Muscle Wasting. Physiol Rev 95, 809–852, doi:10.1152/physrev.00033.2014 (2015).

30 Gramolini, A. O., Belanger, G., Thompson, J. M., Chakkalakal, J. V. & Jasmin, B. J. Increased expression of utrophin in a slow vs. a fast muscle involves posttranscriptional events. Am J Physiol Cell Physiol 281, C1300–1309, doi:10.1152/ajpcell.2001.281.4.C1300 (2001).

31 Mau, T. et al. Mitochondrial energetics in skeletal muscle are associated with leg power and cardiorespiratory fitness in the Study of Muscle, Mobility, and Aging (SOMMA). J Gerontol A Biol Sci Med Sci, doi:10.1093/gerona/glac238 (2022).

32 Gonzalez-Freire, M. et al. Skeletal muscle ex vivo mitochondrial respiration parallels decline in vivo oxidative capacity, cardiorespiratory fitness, and muscle strength: The Baltimore Longitudinal Study of Aging. Aging Cell 17, doi:10.1111/acel.12725 (2018).

33 Bhattacharya, A. et al. Denervation induces cytosolic phospholipase A2-mediated fatty acid hydroperoxide generation by muscle mitochondria. J Biol Chem 284, 46–55, doi:M806311200 [pii] 10.1074/jbc.M806311200 (2009).

34 Triolo, M., Slavin, M., Moradi, N. & Hood, D. A. Time-dependent changes in autophagy, mitophagy and lysosomes in skeletal muscle during denervation-induced disuse. J Physiol 600, 1683–1701, doi:10.1113/JP282173 (2022).

35 Messi, M. L. et al. Resistance Training Enhances Skeletal Muscle Innervation Without Modifying the Number of Satellite Cells or their Myofiber Association in Obese Older Adults. J Gerontol A Biol Sci Med Sci 71, 1273–1280, doi:10.1093/gerona/glv176 (2016).

36 Wang, X. et al. Runx1 prevents wasting, myofibrillar disorganization, and autophagy of skeletal muscle. Genes Dev 19, 1715–1722, doi:10.1101/gad.1318305 (2005).

37 Carreras, D. et al. Epigenetic Changes Governing Scn5a Expression in Denervated Skeletal Muscle. Int J Mol Sci 22, doi:10.3390/ijms22052755 (2021).

38 Balady, G. J. et al. Clinician’s Guide to cardiopulmonary exercise testing in adults: a scientific statement from the American Heart Association. Circulation 122, 191–225, doi:10.1161/CIR.0b013e3181e52e69 (2010).

39 Cawthon, P. M. et al. Strong Relation between Muscle Mass Determined by D3-creatine Dilution, Physical Performance and Incidence of Falls and Mobility Limitations in a Prospective Cohort of Older Men. J Gerontol A Biol Sci Med Sci, doi:10.1093/gerona/gly129 (2018).

40 Linge, J. et al. Body Composition Profiling in the UK Biobank Imaging Study. Obesity (Silver Spring) 26, 1785–1795, doi:10.1002/oby.22210 (2018).

41 Stewart, A. L. et al. CHAMPS physical activity questionnaire for older adults: outcomes for interventions. Med Sci Sports Exerc 33, 1126–1141, doi:10.1097/00005768-200107000-00010 (2001).

42 Kim, D., Paggi, J. M., Park, C., Bennett, C. & Salzberg, S. L. Graph-based genome alignment and genotyping with HISAT2 and HISAT-genotype. Nat Biotechnol 37, 907–915, doi:10.1038/s41587-019-0201-4 (2019).

43 Wells, D. G., McKechnie, B. A., Kelkar, S. & Fallon, J. R. Neurotrophins regulate agrin-induced postsynaptic differentiation. Proceedings of the National Academy of Sciences of the United States of America 96, 1112–1117 (1999).

44 Zhan, W. Z., Mantilla, C. B. & Sieck, G. C. Regulation of neuromuscular transmission by neurotrophins. Sheng li xue bao : [Acta physiologica Sinica] 55, 617–624 (2003).

45 Garcia, N. et al. Localization of brain-derived neurotrophic factor, neurotrophin-4, tropomyosin-related kinase b receptor, and p75 NTR receptor by high-resolution immunohistochemistry on the adult mouse neuromuscular junction. J Peripher Nerv Syst 15, 40–49, doi:10.1111/j.1529-8027.2010.00250.x (2010).

46 Love, M. I., Huber, W. & Anders, S. Moderated estimation of fold change and dispersion for RNA-seq data with DESeq2. Genome Biol 15, 550, doi:10.1186/s13059-014-0550-8 (2014).

