## Supplemental Table 2 for "Higher Expression of Denervation-responsive Genes is Negatively Associated with Muscle Volume and Performance Traits in the Study of Muscle, Mobility and Aging (SOMMA)"

**Supplemental Table 2. Genes with which SOMMA traits had a negative association.**

| Gene Name | Localization | Citations |
|---|---|---|
| CDH15 | A calcium-dependent intercellular adhesion glycoprotein found on muscle fiber side of adult neuromuscular junction | 1. https://pubmed.ncbi.nlm.nih.gov/8921257/ 2. https://pubmed.ncbi.nlm.nih.gov/9638336/ |
| THBS4 | Matricellular protein found at adult neuromuscular junction | 1. https://pubmed.ncbi.nlm.nih.gov/7490284/ |
| CHRND | Delta subunit of acetylcholine receptor found on muscle fiber side of adult neuromuscular junction | 1. https://pubmed.ncbi.nlm.nih.gov/1646821/ 2. https://pubmed.ncbi.nlm.nih.gov/8300573/ |
| CHRNA1 | Alpha subunit of acetylcholine receptor found on muscle fiber side of adult neuromuscular junction | 1. https://pubmed.ncbi.nlm.nih.gov/1646821/ 2. https://pubmed.ncbi.nlm.nih.gov/8300573/ |
| CHRNE | Epsilon subunit of acetylcholine receptor found on muscle fiber side of adult neuromuscular junction | 1. https://pubmed.ncbi.nlm.nih.gov/1646821/ 2. https://pubmed.ncbi.nlm.nih.gov/8300573/ |
| NCAM1 | Glycoprotein found on both muscle fiber and axonal sides of neuromuscular junction | 1. https://pubmed.ncbi.nlm.nih.gov/29175953/ |
| RUNX1 | Transcription factor found in myonuclei | 1. https://pubmed.ncbi.nlm.nih.gov/16024660/ |
| NEFM | A neurofilament protein typically found in axons. Gene has very low baseline expression in skeletal muscle and protein is not detected. | 1. https://www.proteinatlas.org/ENSG00000104722-NEFM/tissue |
| CAV3 | The primary protein structural component of caveolae in skeletal muscle. | 1. https://www.nature.com/articles/ejhg2009103 |
| SCN5A | A sodium channel that is constitutively expressed in adult heart, and although expressed in neonatal skeletal muscle, is typically not expressed in adult skeletal muscle except when denervated. | 1. https://pubmed.ncbi.nlm.nih.gov/2155010/ 2. https://pubmed.ncbi.nlm.nih.gov/1654949/ |
| SCN4A | A sodium channel that is expressed in adult skeletal muscle. | 1. https://pubmed.ncbi.nlm.nih.gov/2155010/ 2. https://pubmed.ncbi.nlm.nih.gov/1654949/ |
| MYOG | A transcription factor that regulates aspects of muscle regeneration, as well as regulating expression of | 1. https://pubmed.ncbi.nlm.nih.gov/20887891/ |

| | | |
|---|---|---|
| | ubiquitin ligases (e.g., atrogin-1/MAFbx) with denervation | |
| GADD45A | Transcription factor that is upregulated in muscle atrophy conditions that include denervation | 1. https://pubmed.ncbi.nlm.nih.gov/22692209/ 2. https://pubmed.ncbi.nlm.nih.gov/23941879/ |
| EFNA2 | Membrane bound protein whose expression increases in injured Schwann cells | 1. https://pubmed.ncbi.nlm.nih.gov/32349983/ |
| CDK5R1 | Receptor for the kinase cdk5 expressed in muscle satellite cells | 1. https://pubmed.ncbi.nlm.nih.gov/28733456/ |
| MUSK | Muscle Specific Kinase found in skeletal muscle membrane and responsible for recruiting proteins to subsynaptic region to anchor acetylcholine receptors to actin cytoskeleton. | 1. https://pubmed.ncbi.nlm.nih.gov/21255125/ 2. https://pubmed.ncbi.nlm.nih.gov/20974278/ |
| RAPSN | A protein that helps anchor acetylcholine receptors to actin cytoskeleton at neuromuscular junction. | 1. https://pubmed.ncbi.nlm.nih.gov/32344108/ |
| KCNC3 | A potassium channel found in various tissues that include skeletal muscle. | 1. https://www.proteinatlas.org/ENSG00000131398-KCNC3/tissue#rna_expression |
| SERPINE2 | Serine protease inhibitor. Gene is expressed in skeletal muscle but protein not detected. | 1. https://www.proteinatlas.org/ENSG00000135919-SERPINE2/tissue |
| NRG1 | Growth factor implicated in regulating acetylcholine receptor number at neuromuscular junction | 1. https://www.ncbi.nlm.nih.gov/pmc/articles/PMC5864756/ |
| P2RX7 | Purinergic receptor involved in formation and function of the neuromuscular junction. | 1. https://www.mdpi.com/1422-0067/24/11/9434 |
| DES | An intermediate filament protein linking contractile proteins to the sarcolemma and organelles. | 1. https://pubmed.ncbi.nlm.nih.gov/33825342/ |
