## Supplemental Table 3 for "Higher Expression of Denervation-responsive Genes is Negatively Associated with Muscle Volume and Performance Traits in the Study of Muscle, Mobility and Aging (SOMMA)"

**Supplemental Table 3. Genes with which SOMMA traits had a positive association.**

| Gene Name | Localization of protein | Citations |
|---|---|---|
| UTRN | A protein structurally and functionally similar to dystrophin expressed primarily in neuromuscular and myotendinous junctions. | 1. https://pubmed.ncbi.nlm.nih.gov/24879867/<br>2. https://www.ncbi.nlm.nih.gov/gene/7402<br>3. https://pubmed.ncbi.nlm.nih.gov/10446253/ |
| F2R | A coagulation factor expressed in osteoclasts and skeletal muscle in response to exercise. | 1. https://pubmed.ncbi.nlm.nih.gov/28377874<br>2. https://pubmed.ncbi.nlm.nih.gov/32226307/ |
| APP | A transmembrane protein expressed at both the pre and post-synaptic sides of the NMJ. | 1. https://pubmed.ncbi.nlm.nih.gov/37175515/<br>2. https://pubmed.ncbi.nlm.nih.gov/15689559/ |
| AKAP1 | A scaffolding protein found in the mitochondria. | 1. https://pubmed.ncbi.nlm.nih.gov/26437602/<br>2. https://pubmed.ncbi.nlm.nih.gov/26610411/ |
| SPOCK1 | A proteoglycan predominately expressed in axons and Schwann cells in adults. Gene expressed in muscle but protein not detected. | 1. https://pubmed.ncbi.nlm.nih.gov/10842087/<br>2. https://www.proteinatlas.org/ENSG00000152377-SPOCK1/tissue |
| MYH9 | A non-muscle myosin found in cytoplasm assisting in cytoskeleton maintenance. | 1. https://pubmed.ncbi.nlm.nih.gov/29679756/ |
| AGRN | Proteoglycan found in the basal lamina of the NMJ in both skeletal muscle and alpha motor neurons. | 1. https://pubmed.ncbi.nlm.nih.gov/28846617/<br>2. https://pubmed.ncbi.nlm.nih.gov/12798796/ |
| SV2A | Synaptic vesicle protein found in the nerve terminals of type I and smaller IIa fibers. Although gene is expressed in skeletal muscle, protein is not detected. | 1. https://pubmed.ncbi.nlm.nih.gov/20843861/<br>2. https://www.proteinatlas.org/ENSG00000159164-SV2A/tissue |
| LRP4 | Agrin receptor found in the skeletal muscle membrane at NMJ. | 1. https://www.ncbi.nlm.nih.gov/pmc/articles/PMC7759742/pdf/fnagi-12-597811.pdf |
| CDK5 | A kinase concentrated at the neuromuscular synapse. | 1. https://pubmed.ncbi.nlm.nih.gov/11276227/ |
| THBS4 | An adhesive glycoprotein that is a member of thrombospondin protein | 1. https://pubmed.ncbi.nlm.nih.gov/37582915/<br>2. https://pubmed.ncbi.nlm.nih.gov/24589453/<br>3. https://pubmed.ncbi.nlm.nih.gov/27669143/ |

| | | |
|---|---|---|
| | family. Stabilizes muscle membranes. | |
| PSEN1 | A protein needed for normal neurogenesis and may have anti-apoptotic activity. | 1. https://www.ncbi.nlm.nih.gov/pmc/articles/PMC9504248/pdf/ijms-23-10970.pdf<br>2. https://www.ncbi.nlm.nih.gov/pmc/articles/PMC10179041/<br>3. https://www.proteinatlas.org/ENSG00000080815-PSEN1/tissue |
| EPHA7 | A juxtacrine signaling receptor concentrated in the post-synaptic membrane of muscle fibers and promotes muscle differentiation. | 1. https://pubmed.ncbi.nlm.nih.gov/11414792/<br>2. https://pubmed.ncbi.nlm.nih.gov/32314958/ |
| KCNC3 | Potassium channel expressed in various CNS neurons, especially purkinje cells, with lower expression in skeletal muscle. | 1. https://pubmed.ncbi.nlm.nih.gov/18448641/<br>2. https://pubmed.ncbi.nlm.nih.gov/34820911/<br>3. https://www.proteinatlas.org/ENSG00000131398-KCNC3/tissue |
| EPHA4 | An ephrin receptor concentrated in the post-synaptic membrane of muscle fibers that is implicated in formation and stability of neuromuscular junction. | 1. https://pubmed.ncbi.nlm.nih.gov/11414792/<br>2. https://pubmed.ncbi.nlm.nih.gov/14729671/ |
| MYH10 | A non-muscle myosin isoform. Gene is expressed in skeletal muscle but protein is not detected. | 1. https://pubmed.ncbi.nlm.nih.gov/22677128/<br>2. https://pubmed.ncbi.nlm.nih.gov/36849436/<br>3. https://www.proteinatlas.org/ENSG00000133026-MYH10/tissue |
